# Binge drinking trajectories across adolescence and early adulthood: Associations with genetic influences for dual-systems impulsive personality traits, alcohol consumption, and alcohol use disorder

**DOI:** 10.1101/2024.10.15.24315471

**Authors:** Alex P. Miller, Kellyn M. Spychala, Wendy S. Slutske, Kim Fromme, Ian R. Gizer

## Abstract

Binge drinking is a relatively common pattern of alcohol use among youth with normative frequency trajectories peaking in emerging and early adulthood. Frequent binge drinking is a critical risk factor for not only the development of alcohol use disorders (AUDs) but also increased odds of alcohol-related injury and death, and thus constitutes a significant public health concern. Changes in binge drinking across development are strongly associated with changes in impulsive personality traits (IPTs) which have been hypothesized as intermediate phenotypes associated with genetic risk for heavy alcohol use and AUD. The current study sought to examine the extent to which longitudinal changes in binge drinking and intoxication frequency across adolescence and early adulthood are associated with genetic influences underlying dual-systems IPTs (i.e., top-down [lack of self-control] and bottom-up [sensation seeking and urgency] constructs) alongside genetic risk for alcohol consumption and AUD. Associations were tested using conditional latent growth curve polygenic score (PGS) models in three independent longitudinal samples (*N*=10,554). Results suggested consistent significant and independent associations across all samples between sensation seeking PGSs and model intercepts (i.e., higher frequency of binge drinking at first measurement occasion) and alcohol consumption PGSs and model slopes (i.e., steeper increases toward peak binge drinking frequency). Urgency PGSs were not significantly associated with changes in binge drinking or intoxication frequency. Collectively, these findings highlight the role of unique but correlated IPT and alcohol-specific genetic factors in the emergence and escalation of binge drinking during adolescence and early adulthood.

---

Binge drinking, characterized by rapid alcohol consumption resulting in elevated blood alcohol concentrations and formally defined as 4+/5+ drinks for women/men on the same occasion (NIAAA, 2015), is particularly prevalent among emerging and young adults. Recent estimates suggest that around 29% of adults aged 18-25 engage in past month binge drinking in the U.S. with nearly 7% doing so five or more times monthly (SAMHSA, 2024). Binge drinking is a well-documented risk factor for subsequent development of alcohol use disorder (AUD) across adolescence and young adulthood (Bonomo et al., 2004), but importantly, also represents a significant societal concern outside of this risk given its proximal link to substantial public health and economic costs (Hingson et al., 2017; Sacks et al., 2015). In the U.S. 13% of deaths attributable to excessive alcohol use are due to acute causes in individuals aged 20 to 34 (CDC, 2022), and binge drinking accounts for more than 75% of the nearly $250 billion economic impact of excessive alcohol use (Sacks et al., 2015).

Normative changes in binge drinking across development have been documented with frequency increasing throughout adolescence and emerging adulthood, peaking during early adulthood, and declining thereafter (Patrick et al., 2019). Correlates of these changes in binge drinking frequency include developmental changes in impulsive personality traits (IPTs; Ashenhurst et al., 2015; McCabe et al., 2021; Quinn et al., 2011; White et al., 2011) consistent with several etiological models describing risky behaviors in adolescence and emerging adulthood (Shulman et al., 2016). For example, dual-systems models emphasize adolescence and emerging adulthood as developmental stages characterized by important changes in the relative contributions of affective-(e.g., urgency) and reward-based drive (e.g., sensation seeking) and self-control on behavior such that inclination towards exciting, pleasurable, and novel activities is not yet fully restrained by a still-developing cognitive control system (Shulman et al., 2016). In this manner, dual-systems models describe impulsive behavior as an expression of bottom-up reward- or emotion-based impulses inadequately inhibited by top-down control (i.e., ‘maturational imbalance’).

In particular, higher levels of sensation seeking and lack of self-control have been shown to be associated with individual differences in binge drinking trajectories across adolescence and early adulthood. For example, there is evidence indicating that higher levels of sensation seeking in early adolescence predict future increases in binge drinking (Drane et al., 2017; Percy et al., 2019) and that changes in both sensation seeking and lack of self-control generally parallel and are bidirectionally associated with changes in binge drinking across adolescence and early adulthood (Ashenhurst et al., 2015; Littlefield et al., 2012; Quinn et al., 2011; White et al., 2011). Additionally, a greater measured imbalance between self-control and sensation seeking in adolescence has been associated with higher levels of binge drinking in emerging adulthood (McCabe et al., 2021). While negative and positive urgency have been shown to relate to measures of binge drinking in cross-sectional analyses (Bø et al., 2016; Lannoy et al., 2017), prior work consistently demonstrates stronger associations with alcohol-related problems than indices of consumption *per se* (Coskunpinar et al., 2013; Stautz & Cooper, 2013), and evidence of longitudinal links between facets of urgency and changes in binge drinking across adolescence or emerging adulthood, more specifically, is limited (Xiao et al., 2009).

Given empirical evidence demonstrating a shared etiology and potential transactional relations between IPTs and binge drinking, behavior genetic studies have attempted to explain the observed interplay between these phenotypes. Twin studies examining the genetic influence and overlap between IPTs and normative and problematic alcohol use have demonstrated that these traits are genetically linked (Ellingson et al., 2013; Khemiri et al., 2016), and while longitudinal investigations of this overlap are largely absent from the twin and molecular genetics literatures, there is evidence to suggest that longitudinal changes in normative and problematic drinking as well as IPTs may be due, in part, to both stable and time-varying genetic influences. As examples from the twin literature, it has been estimated that >80% of individual differences in changes in sensation seeking across adolesence are due to genetic factors (Harden et al., 2012) and genetic influences underlying both alcohol use and AUD may be developmentally variable with adolescent factors being partially distinct from those in adulthood (Edwards & Kendler, 2013; Long et al., 2017). Extending these twin findings, developmentally-informed genome-wide association studies (GWAS) similarly suggest that genetic liability for alcohol use in adolescence may be partially distinct from genetic liability for alcohol use in later developmental periods (Thomas et al., 2023), and GWAS of longitudinal changes in alcohol use and misuse across adolescence and early adulthood have identified common genetic influences associated with steeper developmental trajectories of these alcohol behaviors (Adkins et al., 2015; Edwards et al., 2017).

Though cross-sectional in nature, recent large-scale GWAS of alcohol consumption, AUD, and dual-systems IPT constructs demonstrate shared genetic architecture between IPTs and alcohol use phenotypes (Sanchez-Roige et al., 2023), especially between alcohol consumption and sensation seeking (Miller & Gizer, 2024). Further, though demonstrating appreciable overlap in their findings, recent GWAS of alcohol consumption and AUD have consistently revealed specific and separable genetic influences on these traits (Mallard et al., 2022). In the post-GWAS era, variant-level summary statistics from GWAS such as these can be used to calculate polygenic scores (PGSs) in independent samples as a means of estimating individual genetic liability for a specific trait relative to a population. Few studies to date have used IPT PGSs to predict alcohol use phenotypes (e.g., risk taking PGSs; Karlsson Linnér et al., 2019; Ksinan et al., 2019) and even fewer have examined the longitudinal impact of these PGSs on alcohol use phenotypes. However, the developmental relevance of the genetic overlap between these traits has been indirectly supported in mediation studies showing that sensation seeking partially mediates associations between alcohol dependence PGSs and subsequent alcohol problems in adolescence (Li et al., 2017) and alcohol problems PGSs and subsequent binge drinking in emerging adulthood (Lannoy et al., 2023).

The converging lines of evidence reviewed here highlight the need for research focusing on well-defined components of IPTs and specific alcohol use behaviors that are informed by developmental models of adolescent and young adult alcohol use progression. Consistent with this viewpoint, the present study sought to integrate research from the fields of developmental psychopathology, addiction science, and human genetics with the goal of examining the relative contributions of PGSs for dual-systems IPTs (i.e., top-down [lack of self-control] and bottom-up [sensation seeking and urgency] constructs; Miller & Gizer, 2023), and alcohol consumption and AUD (Miller & Gizer, 2024) to change in binge and intoxication frequency across adolescence and early adulthood using conditional latent growth curve modelling in three independent longitudinal samples: The National Longitudinal Study of Adolescent to Adult Health (Add Health), the Genes and New Experiences Study (GENES), and the Avon Longitudinal Study of Parents and Children (ALSPAC). Given existing knowledge about the longitudinal phenotypic relations between IPTs and binge drinking across adolescence and early adulthood, research highlighting the developmental relevance of dual-systems IPT models, and specificity of genetic effects for alcohol consumption versus AUD, it was hypothesized that (1) IPT PGSs, especially sensation seeking and lack of self-control, would show significant associations with changes in binge and intoxication frequency; (2) alcohol consumption PGSs would predict changes in binge and intoxication frequency more strongly than AUD PGSs; and (3) IPT and alcohol consumption PGSs would demonstrate partially unique prediction of changes in binge and intoxication frequency. While there is evidence to suggest that negative and positive urgency are associated with binge drinking cross-sectionally, thus motivating their inclusion in the present study, less is known regarding the impact of urgency traits on binge drinking trajectories, and as such, these analyses were conducted without *a priori* hypotheses.

## Methods

### Discovery Samples

GWAS summary statistics of dual-systems IPTs (lack of self-control, sensation seeking, and urgency), alcohol consumption, and AUD (Miller & Gizer, 2023, 2024) were utilized to generate PGSs in longitudinal target samples. GWAS summary statistics for dual-systems IPTs included GenomicSEM (Grotzinger et al., 2019) factors for sensation seeking (*N*^^^=710,791), lack of self-control (*N*^^^=27,656), and urgency (*N*^^^=28,316). Summary statistics for alcohol traits included a GenomicSEM alcohol consumption factor (*N*^^^=1,388,120) and an AUD GWAS meta-analysis (*n_effective_*=220,182). All GWAS samples were restricted to individuals of European ancestry for greater generalizability to longitudinal target samples which were also restricted to individuals of European ancestry. See Supplementary Methods for descriptions of genotyping, imputation, and quality control for target samples and previous papers for extended descriptions of discovery GWAS (Miller & Gizer, 2023, 2024).

### Target Samples

The current study utilized data from unrelated participants who (1) completed one or more assessments of binge or intoxication frequency across available timepoints (i.e., pairwise deletion) and (2) for whom genotype data were available. Descriptive statistics for all samples are included in Supplementary Tables 1-4. Proportions of endorsed binge and intoxication frequency responses are shown in Figures 1A, 2A, and 3A. Secondary analyses of all data were considered exempt by the University of Missouri Institutional Review Board.

**Figure 1.**
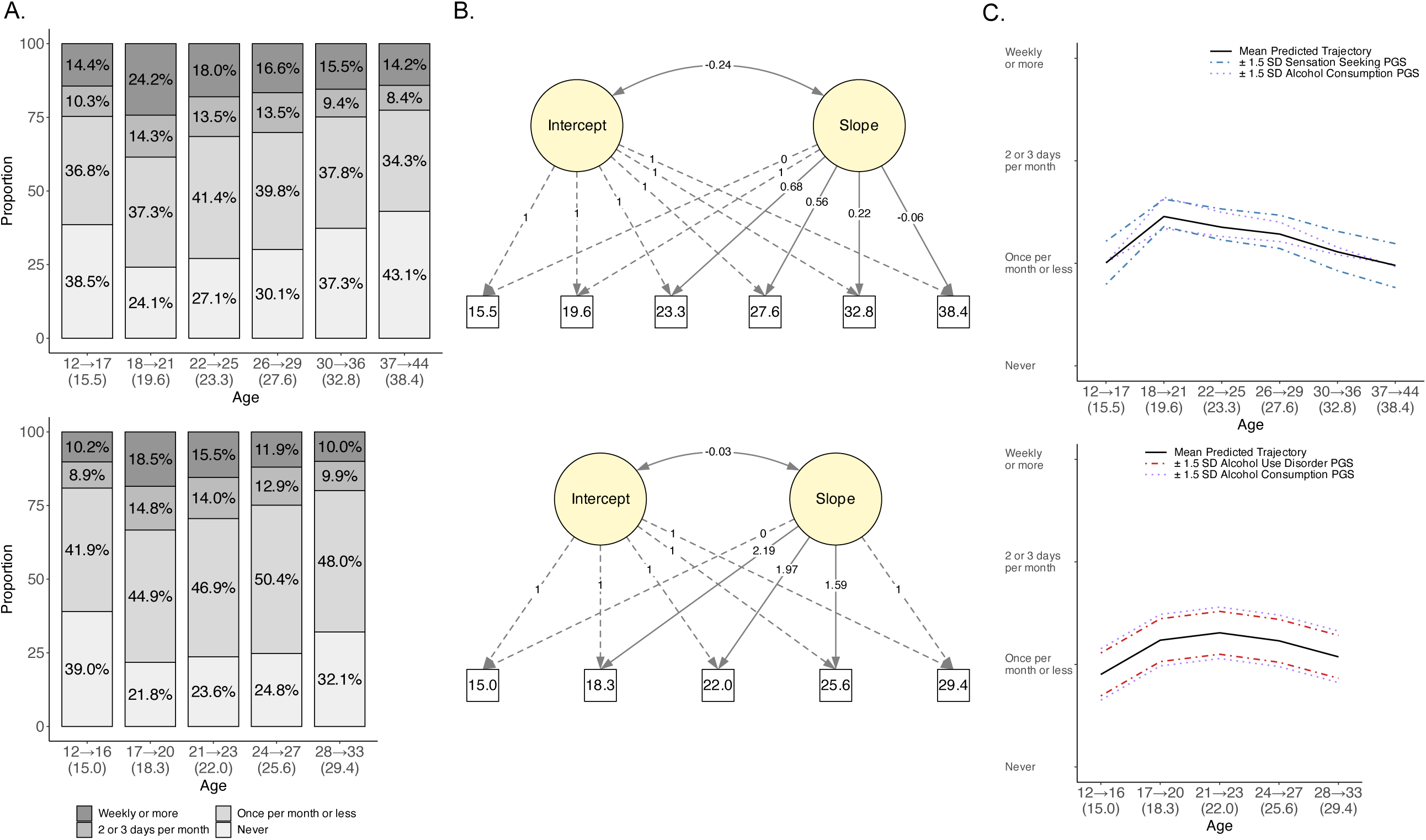
Latent growth curve modeling of binge (*n*=4,710) and intoxication (*n*=4,538) frequency change and associations with polygenic scores in Add Health. **(A)** Proportions of responses to binge frequency (top) and intoxication frequency (bottom) questions across age groups. Values reported on *X*-axis reflect ranges of ages for each age group with mean age in parentheses. **(B)** Path diagrams of unconditional free curve slope intercept models for binge frequency (top) and intoxication frequency (bottom). Dashed lines indicate fixed parameters, and solid lines indicate freely estimated unstandardized parameter estimates. **(C)** Plotted trajectories based on parameter estimates from conditional free curve slope intercept PGS model for binge frequency (top) and intoxication frequency (bottom) displaying PGSs with significant associations (*P* < .05). Lines depict the mean predicted trajectory and trajectories for high and low PGSs (±1.5 standard deviation around the mean).

### The National Longitudinal Study of Adolescent to Adult Health (Add Health)

Add Health is a longitudinal study of a nationally-representative sample of adolescents born between March 1974 and July 1983 in the United states and first recruited for participation in grades 7-12 in 1994-1995 (Harris et al., 2019). Currently, data from five assessment waves have been collected (Wave 1: 1994-1995, Wave 2: 1996, Wave 3: 2001-2002, Wave 4: 2008, Wave 5: 2016-2018). Longitudinal assessments of binge drinking frequency (“During the past 12 months, on how many days did you drink {five=male/four=female} or more drinks in a row?”) assessed at all five waves and intoxication frequency (“During the past 12 months, on how many days have you been drunk or very high on alcohol?”) assessed at Waves 1-4 served as the primary phenotypes of interest.

As Add Health follows a cohort-sequential design where individuals across an age range are assessed at each wave, data were restructured to provide age-based measurement and collapsed into age groups to maximize coverage for longitudinal analyses and correspondence with lifespan development stages. Given that Wave 1 and Wave 2 were approximately 1 year apart, some individuals were interviewed at the same age at Waves 1 and 2 (*n=*1,657), and at later assessments, some individuals were assessed more than once in a given age group (e.g., age 14 years at Wave 1, age 16 years at Wave 2, thus both assessments were collapsed into the age 12→17 group). In such instances, if individuals provided binge frequency or intoxication frequency data at only one wave, their response was taken from that wave. If individuals provided data at both waves, their response was taken from the later wave/age. The final samples included 4,710 individuals (46.9% male) with valid responses for binge frequency and 4,538 individuals (47.2% male) with valid responses for intoxication frequency.

### Genes and New Experiences Study (GENES)

GENES is an extension of The UT Experience! study for which a cohort of college freshman, born between September 1985 and January 1986, were recruited in 2004 to participate in a longitudinal study investigating individual differences in a range of psychological outcomes.

Assessments began the summer prior to freshman year and continued every semester for seven semesters, followed by two additional yearly assessments, and a final follow-up assessment three to seven years later (average ages=18.4, 18.8, 19.2, 19.8, 20.2, 20.8, 21.2, 21.8, 22.8, 23.8, and 27.5 years old; see Ashenhurst et al., 2015 for further details). Longitudinal assessments of binge drinking (“During the last 3 months, how many times did you have {five=male/four=female} or more drinks at a sitting?”) and intoxication frequency (“During the last 3 months, how many times did you get drunk (not just a little high) on alcohol?”) served as primary phenotypes. The final sample included 371 individuals (35.8% male) with valid responses for binge and intoxication frequency.

### Avon Longitudinal Study of Parents and Children (ALSPAC)

ALSPAC is a multi-generation, longitudinal birth cohort study conducted at the University of Bristol in South West UK and has followed 14,451 mothers recruited during pregnancy, along with their partners and their offspring born between 1991 and 1992 in Avon, UK (Boyd et al., 2013; Fraser et al., 2013; Northstone et al., 2019). Ethical approval for the study was obtained from the ALSPAC Ethics and Law Committee and the Local Research Ethics Committees. The ALSPAC website contains details of all data that are available through a fully searchable data dictionary and variable search tool (http://www.bristol.ac.uk/alspac/researchers/our-data/). The current study focused on ALSPAC offspring, and the final sample included 5,473 individuals (43.5% male) with valid binge frequency (“How often, during the past year, have you had six or more units [standard drinks] on one occasion?”) responses at a minimum of one of the seven timepoints examined (target ages=16, 17.5, 18, 20, 22, 24, and 28 years old). Of note, as a UK-based sample, ALSPAC study definitions of standard drinks (http://www.bris.ac.uk/media-library/sites/alspac/documents/participants/drinkogram.pdf) differ slightly from those used in US-based samples (8g of alcohol in ALSPAC *vs*. 14g of alcohol in Add Health and GENES). Thus, whereas a binge episode in Add Health and GENES equates to roughly 70g of alcohol, a binge episode in the ALSPAC sample is closer to 48g of alcohol.

### Polygenic Scores

PGSs were generated using discovery sample GWAS summary statistics for sensation seeking, lack of self-control, urgency, alcohol consumption, and AUD in each target sample. To avoid overlap between discovery samples and target samples, alternate GWAS summary statistics for PGS construction were computed for alcohol consumption and AUD for ALSPAC and Add Health, respectively (see Supplementary Methods for additional details on alternate GWAS summary statistics).

Weights for PGSs were generated using SBayesR (Lloyd-Jones et al., 2019), which employs a Bayesian framework to shrink single nucleotide polymorphism (SNP) effect sizes while maximizing variance explained and accounting for linkage disequilibrium to generate high probability distributions of SNP effects. Individual PGSs were then scored in PLINK 1.9 (Chang et al., 2015) using the SBayesR-derived weights. See Supplementary Methods for additional details regarding SBayesR implementation and Supplementary Tables 5-8 for numbers of SNPs used to generate PGSs and correlations between respective PGSs in each sample.

### Data Analysis

Binge and intoxication frequency response data were coded as follows to generate similar scales for each sample: Add Health, 1=*Never*, 2=*Once per month or less*, 3=*2 or 3 days per month*, 4=*Weekly or more*; GENES and ALSPAC, 1=*Never*, 2=*Less than monthly*, 3=*Monthly*, 4=*Weekly or more* (see Supplementary Methods for additional details). Longitudinal changes in binge and intoxication frequency were modeled in each target sample using unconditional latent growth curve models (LGCMs) to estimate developmental trends in binge drinking using random effects, conceptualized as latent growth factors. Five models were fit for each of the available binge/intoxication measures in each sample: (1) intercept only, (2) intercept and linear slope, (3) intercept and partly nonlinear slope (quadratic), (4) intercept and piecewise linear slopes, and (5) intercept and fully nonlinear slope (i.e., latent basis or free curve slope intercept [FCSI]; (Meredith & Tisak, 1990). For all models containing linear slope estimates, loadings were scaled to reflect actual time between assessments with T1 fixed to 0 and all subsequent timepoints reflecting time elapsed since T1 in years divided by 10. For all FCSI models, except those for Add Health binge frequency, the first and last slope factor loadings were fixed at 0 and 1, respectively, with the interpretation that all subsequent change is relative to the total change that occurred over all timepoints. For Add Health, mean binge frequency at the final timepoint was lower than at the first timepoint. To obtain more interpretable loadings, the first and second slope factor loadings were fixed at 0 and 1 instead of the first and final loadings, with the interpretation that all subsequent change is relative to that which occurred between the first two timepoints.

Within each growth function specification, two sub-models were specified: unstructured vs. structured residuals (LGCM-SR; Curran et al., 2014). LGCM-SR models included autoregressive, time-specific residual latent factors that account for deviations between intercept and slope implied trajectories by modeling associations between adjacent timepoints not accounted for by growth factors.

All LGCMs were fit in *lavaan* (v0.6-8; Rosseel, 2012) in R (R Core Team, 2023) using weighted least squares with mean and variance adjustment estimation with θ parameterization (probit link), which equates the scale of the underlying distribution across timepoints to allow for the examination of continuous changes in means and variances over time. This estimation approach provides robust standard errors, does not assume normally distributed variables, and is commonly used for modelling categorical or ordered data (Brown, 2015). Model fit comparisons were conducted using appropriate robust versions of common fit indices concurrently (Brosseau-Liard et al., 2012; Brosseau-Liard & Savalei, 2014), including the *χ*^2^_S_ fit statistic, the comparative fit index (CFI_S_), the Tucker-Lewis index (TLI_S_), the root mean square error of approximation (RMSEA_S_), and the standardized root mean square residual (SRMR). CFI_S_ and TLI_S_ values ≥.95, and RMSEA values ≤.06 were used to indicate good model fit. SRMR values <.10 and <.05 were used to indicate acceptable and good model fit, respectively. For nested LGCMs (unstructured vs. structured residuals), the scaled *χ*^2^ difference test for weighted least squares models (Satorra, 2000) was used to assess the significance of change in model fit provided by structuring residuals.

Following selection of the best-fitting unconditional LGCMs, conditional models were fit in which growth factors were regressed on all five PGS simultaneously to directly compare predictive performance of each PGS when controlling for all others. Covariates included sex and the first 10 genomic ancestry principal components (PCs) in each sample to control for possible population stratification. All PGSs and PCs were standardized (*M=*0, *SD=*1) prior to analysis.

Changes in growth factor variance explained (Δ*R*^2^) by each significant PGS predictor, controlling for all other PGSs and covariates, were estimated using an approach described by Hayes (2021). Model code is available upon request to the first author. Analyses were not preregistered and should be considered exploratory.

## Results

Fit statistics for best-fitting unconditional LGCMs for each target sample and phenotype are shown in Table 1 and path diagrams are depicted in Figures 1B, 2B, and 3B. Standardized regression coefficients and 95% CIs from PGS conditional LGCMs can be found in Table 2. Figures 1C, 2C, and 3C depict model-implied mean growth trajectories compared to growth trajectories for individuals with high or low scores (i.e., ±1.5 *SD* around the mean) for significant PGSs.

**Table 1.** Model fit statistics for best-fit unconditional latent growth curve models.

| Sample | Phenotype | Model | $\chi^2_s$ | *df* | *P* | RMSEAs [90% CI] | SRMR | CFIs | TLIs |
| --- | --- | --- | --- | --- | --- | --- | --- | --- | --- |
| Add Health | Binge Frequency | FCSI | 169.738 | 15 | < .001 | 0.047 [0.041, 0.053] | 0.084 | 0.930 | 0.930 |
| Add Health | Intoxication Frequency | FCSI | 76.201 | 9 | < .001 | 0.041 [0.032, 0.049] | 0.069 | 0.928 | 0.920 |
| GENES | Binge Frequency | FCSI-SR | 106.589 | 50 | < .001 | 0.055 [0.041, 0.070] | 0.028 | 0.994 | 0.993 |
| GENES | Intoxication Frequency | FCSI-SR | 85.153 | 50 | .001 | 0.044 [0.027, 0.059] | 0.027 | 0.995 | 0.994 |
| ALSPAC | Binge Frequency | Piecewise-SR (Knot at T4) | 241.019 | 18 | < .001 | 0.048 [0.042, 0.053] | 0.023 | 0.984 | 0.981 |

*Note:* FCSI = free curve slope intercept; RMSEAs = root mean square error of approximation; SRMR = standardized root mean squared residual; CFI = comparative fit index; TLI = Tucker-Lewis index; -SR models include an autoregressive residual structure. All fit indices other than the SRMR are adjusted for non-normality of data (i.e., categorical outcomes).

**Table 2.**
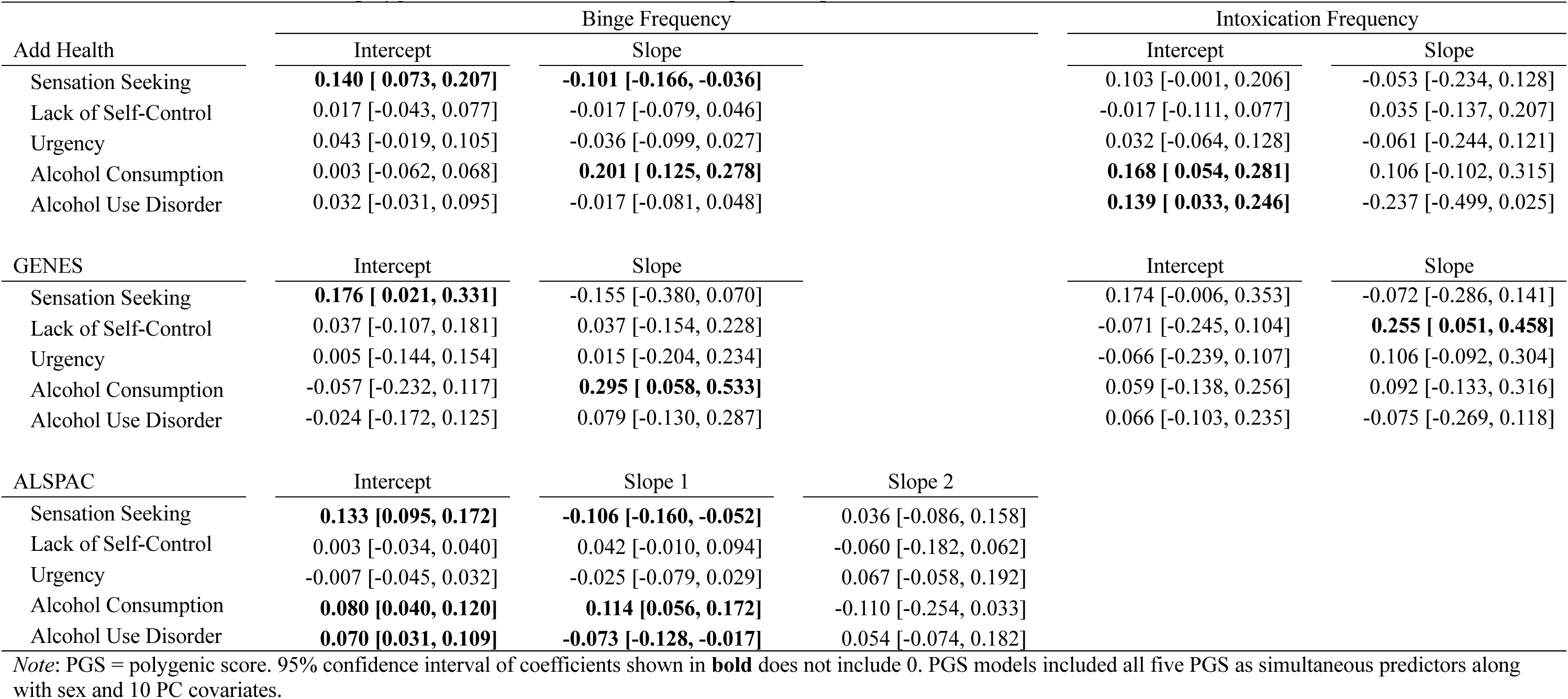
Standardized coefficients of polygenic score associations with intercept and slope factors from conditional LGCMs.

Across samples, there was a general trend for binge and intoxication frequency to increase across adolescence (age 12-18 years), peak in emerging young adulthood (age ∼21 years), and then decline across adulthood. Participants in the GENES sample, however, showed a deviation from this pattern as binge and intoxication frequency showed an increase at the last timepoint (age ∼27 years) relative to the previous wave (age ∼24 years). Correlations between adjacent age groups were modest in the Add Health sample (binge frequency–*r*=.21-.50, intoxication frequency–*r*=.12-.41) and higher in the ALSPAC (binge frequency–*r*=.43-.60) and GENES (binge frequency–*r*=.58-.77, intoxication frequency–*r*=.58-.76) samples (Supplementary Tables 1-4). The high autocorrelations in the GENES sample were expected given the shorter time intervals (∼3 months) between assessments from T1-T8.

### Unconditional Models

#### The National Longitudinal Study of Adolescent to Adult Health (Add Health)

The best-fitting unconditional LGCMs for binge and intoxication frequency in the Add Health sample were FCSI models with unstructured residuals (see Figure 1B for path diagrams and Supplementary Tables 9-10 for fit statistics). Both FCSI models provided adequate-to-good fit to the data, as evidenced by model fit indices (Table 1) and parameter estimates (Supplementary Tables 11-12), which were consistent with observed trajectories described above. All variance and covariance estimates for growth factors were statistically significant in the binge frequency model (*ζ*_I=_0.67, *P=*2.14×10^-42^; *ζ*_S=_0.38, *P=*.0004; *ψ*_I-S=_-0.21, *P=*.001), while only the intercept factor was statistically significant in the intoxication frequency model (*ζ*_I=_0.49, *P=*.001; *ζ*_S_=0.11, *P*=.061; *ψ*_I-S=_-0.04, *P=*.647).

#### Genes and New Experiences Study (GENES)

In the GENES sample, the best-fitting unconditional LGCMs for binge and intoxication frequency were again FCSI models, though in this sample structured residuals were included (LGCM-SR; see Figure 2B for path diagrams and Supplementary Tables 13-14 for fit statistics of all models tested). These models provided good fit to the data, as evidenced by model fit indices (Table 1) and parameter estimates (Supplementary Tables 15-16) and were consistent with observed trajectories. All variance and covariance estimates for growth factors were statistically significant in the binge frequency model (*ζ*_I=_3.06, *P=*1.11×10^-12^; *ζ*_S=_1.58, *P=*.005; *ψ*_I-S=_-1.41, *P=*.012), while only the intercept factor was statistically significant in the intoxication frequency model (*ζ*_I=_1.42, *P=*.002; *ζ*_S_=0.81, *P*=.064; *ψ*_I-S=_-0.25, *P=*.568).

**Figure 2.**
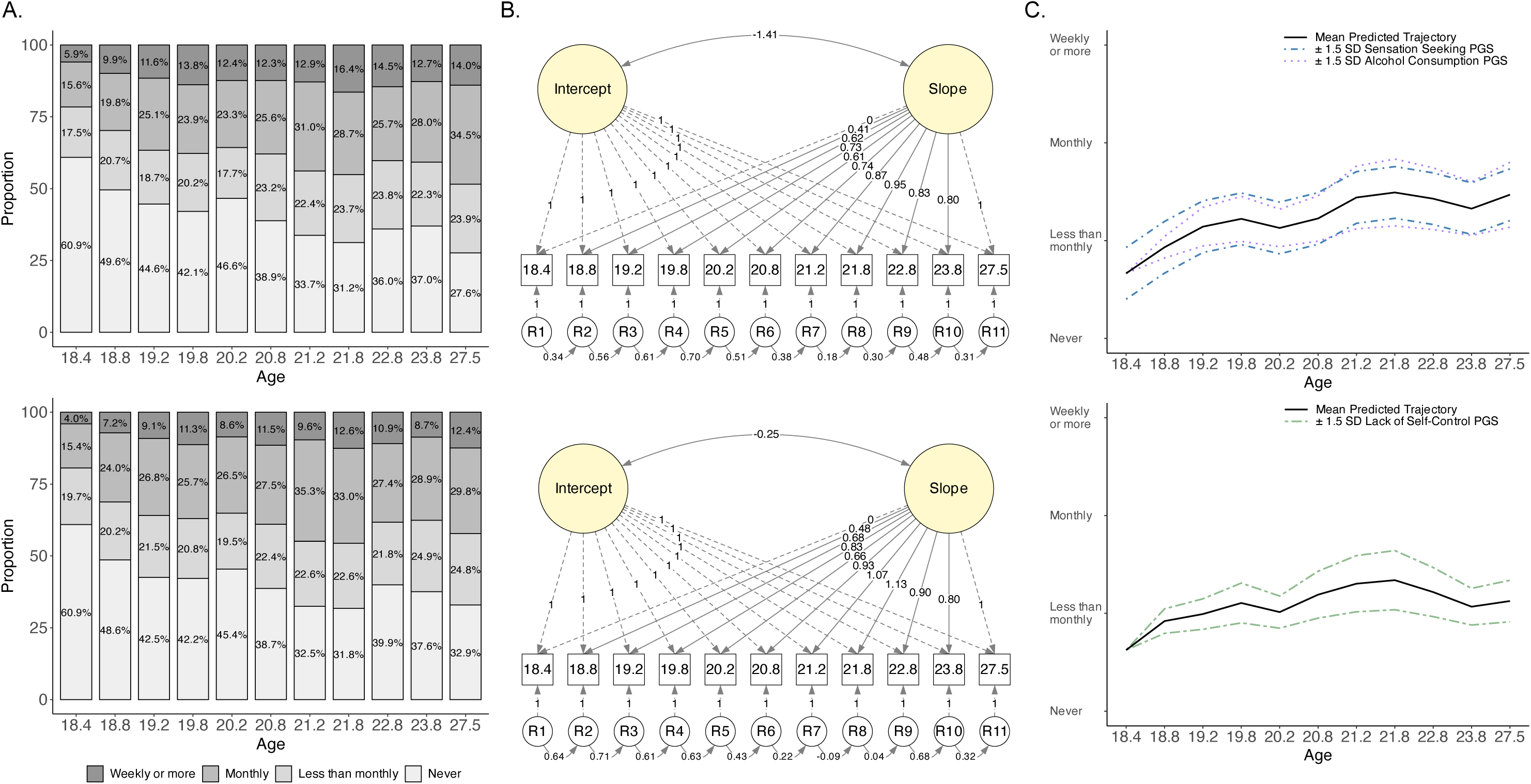
Latent growth curve modeling of binge and intoxication frequency change (*n*=371) and associations with polygenic scores in GENES. **(A)** Proportions of responses to binge frequency (top) and intoxication frequency (bottom) questions across timepoints. Values reported on *X*-axis reflect mean age at each assessment. **(B)** Path diagrams of unconditional free curve slope intercept models with structured residuals for binge frequency (top) and intoxication frequency (bottom). Dashed lines indicate fixed parameters, and solid lines indicate freely estimated unstandardized parameter estimates. **(C)** Plotted trajectories based on parameter estimates from conditional free curve slope intercept PGS model with structured residuals for binge frequency (top) and intoxication frequency (bottom) displaying PGSs with significant associations (*P* < .05). Lines depict the mean predicted trajectory and trajectories for high and low PGSs (±1.5 standard deviation around the mean).

#### Avon Longitudinal Study of Parents and Children (ALSPAC)

The best-fitting unconditional LGCM for binge frequency in the ALSPAC sample was the piecewise model with structured residuals (see Figure 3B for path diagram and Supplementary Table 17 for fit statistics of all models tested). Given the obvious peak in binge frequency growth trends, the ‘knot’ for this model was placed at T4 with T1-T4 representing one linear growth trajectory and T4-T7 representing a second. This model provided good fit to the data, as evidenced by model fit indices (Table 1) and parameter estimates (Supplementary Table 18) and were consistent with observed trajectories. Variance and covariance estimates for intercept and slope 1 were statistically significant (*ζ*_I=_1.32, *P=*2.08×10^-67^; *ζ*_S1=_7.02, *P<*1×10^-300^; *ψ*_I-S1=_-1.79, *P=*6.10×10^-48^) as was the covariance between slope 1 and slope 2 (*ψ*_S1-S2=_-0.46, *P=*.004). The variance of slope 2 and the covariance between the intercept and slope 2 were not significant (*ζ*_S2_=0.16, *P*=.079; *ψ*_I-S2=_-0.05, *P=*.199).

**Figure 3.**
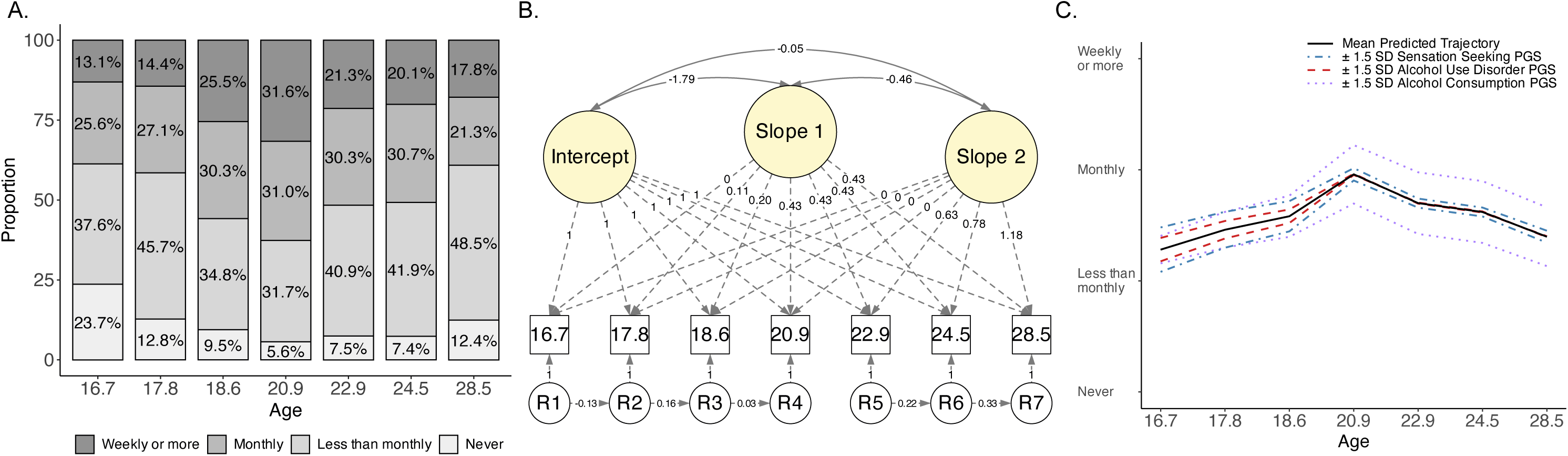
Latent growth curve modeling of binge frequency changes (*n*=5,473) and associations with polygenic scores in ALSPAC. **(A)** Proportions of responses to binge frequency questions across timepoints. Values reported on *X*-axis reflect mean age at each assessment. **(B)** Path diagrams of unconditional piecewise latent growth curve models with structured residuals for binge frequency. Dashed lines indicate fixed parameters, and solid lines indicate freely estimated unstandardized parameter estimates. **(C)** Plotted trajectories based on parameter estimates from conditional piecewise latent growth curve PGS model with structured residuals for binge frequency displaying PGSs with significant associations (*P* < .05). Lines depict the mean predicted trajectory and trajectories for high and low PGSs (±1.5 standard deviation around the mean).

### Conditional Models

#### The National Longitudinal Study of Adolescent to Adult Health (Add Health)

For the Add Health sample, the binge and intoxication frequency PGS models showed multiple significant associations with both intercept and slope factors (Table 2; Figure 1C). Specifically, sensation seeking PGSs (*β=*0.14, *SE*=0.03, *P*<.001, Δ*R*^2^=.002) were positively associated with the *binge frequency* intercept factor. Alcohol consumption PGSs (*β=*0.17, *SE*=0.06, *P*=.004, Δ*R*^2^=.008) and AUD PGSs (*β=*0.14, *SE*=0.05, *P*=.010, Δ*R*^2^=.019) were positively associated with the *intoxication frequency* intercept factor, together explaining 3.45% of the variance in this factor.

Sensation seeking PGSs were negatively associated with the *binge frequency* slope factor (*β=*-0.10, *SE*=0.03, *P*=.002, Δ*R*^2^=.0003) such that PGSs 1.5 *SD* above the mean resulted in Δ*β*=−0.04 from T1-T2 (modeled peak) and Δ*β*=+0.02 from T2-T5 relative to the mean trajectory. In contrast, alcohol consumption PGSs were positively associated with the *binge frequency* slope factor (*β=*0.20, *SE*=0.04, *P*<.001, Δ*R*^2^=.008) such that PGSs 1.5 *SD* above the mean resulted in Δ*β*=+0.19 from T1-T2 and Δ*β*=−0.10 from T2-T5 relative to the mean trajectory. Together these PGSs explained 0.79% of *binge frequency* slope factor variance. No PGSs were associated with the *intoxication frequency* slope factor.

#### Genes and New Experiences Study (GENES)

For the GENES sample, sensation seeking PGSs were positively associated with the intercept factor in the *binge frequency* model (*β=*0.18, *SE*=0.08, *P*=.026, Δ*R*^2^=.024), but no PGSs were associated with the intercept factor in the *intoxication frequency* model (Table 2; Figure 2C). When looking at the slope factors, alcohol consumption PGSs were significantly associated with the *binge frequency* slope factor (*β=*0.30, *SE*=0.12, *P*=.014, Δ*R*^2^=.054) such that PGSs 1.5 *SD* above the mean resulted in Δ*β*=+0.23 from T1-T11 relative to the mean trajectory, and lack of self-control PGSs were significantly associated with the *intoxication frequency* slope factor (*β=*0.25, *SE*=0.10, *P*=.014, Δ*R*^2^=0.077) such that PGSs 1.5 *SD* above the mean resulted in Δ*β*=+0.19 from T1-T11 relative to the mean trajectory.

#### Avon Longitudinal Study of Parents and Children (ALSPAC)

For ALSPAC, only *binge frequency* data were available. Three PGSs were positively associated with the *binge frequency* intercept factor: sensation seeking (*β*=0.13, *SE*=0.02, *P*<.001, Δ*R*^2^=.016), alcohol consumption (*β*=0.08, *SE*=0.02, *P*<.001, Δ*R*^2^=.006), and AUD (*β*=0.07, *SE*=0.02, *P*<.001, Δ*R*^2^=.004; Table 2; Figure 3C). Collectively, these three PGS explained 3.49% of *binge frequency* intercept factor variance. The same three PGS were also associated with the *binge frequency* slope 1 factor: sensation seeking (*β*=-0.11, *SE*=0.03, *P*<.001, Δ*R*^2^=.009), alcohol consumption (*β*=0.11, *SE*=0.03, *P*<.001, Δ*R*^2^=.012), and AUD (*β*=-0.07, *SE*=0.03, *P*=.010, Δ*R*^2^=.005). Collectively, these three PGS explained 2.10% of *binge frequency* slope 1 factor variance. Alcohol consumption PGSs 1.5 *SD* above the mean resulted in Δ*β*=+0.08 from T1-T4 (modeled peak) relative to the mean trajectory, while sensation seeking and AUD PGSs 1.5 *SD* above the mean resulted in Δ*β*=−0.08 and Δ*β*=−0.05 from T1-T4 relative to the mean trajectory, respectively. No PGSs were associated with the second *binge frequency* slope factor.

## Discussion

Using data from three independent longitudinal samples, the current study provides initial evidence that genetic influences for dual-systems IPTs, alcohol consumption, and AUD exhibit associations with changes in binge drinking and intoxication across adolescence and early adulthood. Multivariable conditional LGCMs revealed distinct roles for sensation seeking, lack of self-control, alcohol consumption, and AUD PGSs in the prediction of binge and intoxication frequency changes. Consistent patterns of associations were observed across samples: (1) sensation seeking PGSs were positively associated with binge frequency model intercepts; (2) alcohol consumption PGSs were positively associated with binge frequency model slopes; and (3) urgency PGSs were not associated with any modeled growth parameters. Within each sample and phenotype, more specific findings were observed for lack of self-control PGSs (positive association with intoxication slope factor in GENES) and AUD PGSs (positive associations with intoxication intercept in Add Health and binge intercept in ALSPAC and negative associations with initial binge slope in ALSPAC). The results of the current study are discussed first in the context of replicable patterns of PGS associations across samples, followed by unique sample-specific associations, and finally a general discussion of findings, limitations, and future directions.

### Findings Across Samples

In each sample, conditional LGCMs demonstrated a positive association between sensation seeking PGSs and model intercepts of binge frequency controlling for other tested PGSs and covariates. Additionally, observed relations between sensation seeking PGSs and changes in binge drinking frequency in Add Health and ALSPAC were negative, albeit small, such that those with higher PGSs demonstrated slower increases leading up to peak binge frequency. Together, these results suggest that higher genetic propensity for sensation seeking is associated with higher binge drinking frequencies in early and adolescence independent of genetic risk for other IPT or alcohol use phenotypes. Further, these genetic influences may lead to an earlier onset of binge drinking that occurs outside the developmental timeframes captured by the included studies (i.e., prior to age 16) that then results in a protracted or more gradual increase in binge drinking up to the modeled peak.

These findings are broadly consistent with phenotypic studies demonstrating that sensation seeking is robustly associated with adolescent alcohol use and binge drinking more specifically (Stautz & Cooper, 2013). Longitudinal studies focusing on early adolescence and childhood suggest that sensation seeking predicts earlier onset and more persistent alcohol use (Malmberg et al., 2010; Peeters et al., 2014) as well as increases in binge drinking frequency (Drane et al., 2017). These findings have led several researchers to hypothesize that a primary contribution of sensation seeking to alcohol use and alcohol use trajectories is a direct relation to earlier experimentation with alcohol, which has been demonstrated in multiple studies (Cappelli et al., 2020; Jensen et al., 2017), including those focused on alcohol sipping as the earliest form of experimentation (Jackson et al., 2015; Watts et al., 2023), which then produces subsequent increases in riskier forms of alcohol use such as binge drinking (Doumas et al., 2019). The present findings further support this hypothesis and suggest that longitudinal studies beginning in childhood (e.g., the Adolescent Brain Cognitive Development (ABCD) Study; Volkow et al., 2018) will be necessary to more fully study the relations of sensation seeking and its genetic underpinnings on binge drinking trajectories.

A second finding across samples was that alcohol consumption PGSs were consistently associated with binge frequency slope variances such that those with higher genetic loading for increased alcohol consumption exhibited steeper increases to peak binge frequency. This result replicates those of prior studies suggesting genetic predispositions to higher alcohol consumption predict greater levels of both typical and binge alcohol use during emerging adulthood (Ksinan et al., 2019; Lannoy et al., 2023) as well as increases in regular alcohol use quantity over a similar developmental span (Deak et al., 2022). Interestingly, in Add Health, higher alcohol consumption PGSs were associated, not only with steeper increases in binge drinking frequency from adolescence to peak in emerging adulthood as was found in GENES and ALSPAC, but also steeper declines in binge frequency from emerging adulthood to early middle adulthood.

Similar to the sensation seeking findings, it is important to consider the developmental timeframes covered by the individual studies when interpreting these results. The assessment of binge frequency extended the furthest into adulthood in Add Health relative to the other samples by roughly a decade. While the alcohol consumption PGS predicted a swifter increase in binge drinking frequency during the transition out of adolescence relative to the mean, these same genetic influences did not demonstrate a tendency to delay ‘maturing out’ of heavier alcohol consumption, and in fact, appeared to increase the rate of reductions in binge drinking frequency. This result suggests that other genetic influences, not captured by the alcohol consumption PGS, and possible unexamined environmental variables, such as relevant role transitions (e.g., lack of employment, changes in marital status; Lee & Sher, 2018), may influence delays in this binge drinking desisting process. Thus, the alcohol consumption PGSs used in the current study may primarily index risk for early escalation with additional genetic risk factors conferring risk for continued escalation in early adulthood (Edwards & Kendler, 2013; Thomas et al., 2023). Moreover, these findings extend previous literature by showing that alcohol consumption PGSs reliably predict increases in episodic heavy drinking independent of other plausible genetic risk factors (e.g., AUD PGSs).

To put these findings into the context of proposed models of AUD (e.g., Koob & Volkow, 2016), the present findings suggest that both the sensation seeking and alcohol consumption PGSs are particularly relevant to the early stages of drinking that are driven by the positively reinforcing effects of alcohol rather than later stages that are driven more by relief from alcohol’s negative effects. Notably, these influences may operate through multiple direct and indirect pathways. For example, multiple studies have demonstrated a potential pathway through which the impact of genetic risk for alcohol consumption on elevated levels of alcohol use is partially mediated by higher levels of sensation seeking (Lannoy et al., 2023; Su et al., 2021), highlighting the shared genetic architecture between alcohol consumption and sensation seeking (Miller & Gizer, 2024). The present study provides evidence of the unique, direct effects of the genetic influences underlying sensation seeking and alcohol consumption on changes in binge drinking and drinking to intoxication. Nonetheless, other variables are likely to be relevant to these relations. For example, sensation seeking may be important for early experimentation with alcohol, but there is also evidence that sensation seeking may be related to the subjective response to alcohol and the stimulating effects in particular (Ray et al., 2006; Scott & Corbin, 2014). Genetic effects may also operate through correlations with environmental variables, such as exposure to heavy drinking peers. Previous studies have suggested that those higher in genetic risk for heavy alcohol use and other externalizing behavior self-select into peer groups engaging in these behaviors (gene-environment correlation) but are also more susceptible to the influences of these peers (gene-environment interaction) as well (Harden et al., 2008).

Finally, it is worth noting that urgency PGSs were not significant predictors of outcomes in any of the conducted analyses. This may reflect both a lack of power inherent in the PGSs calculated using summary statistics from the urgency GWAS (Miller & Gizer, 2023) and the potentially weaker genetic influences underlying the smaller observed phenotypic associations between urgency and binge drinking or alcohol consumption more broadly. To the first point, while the urgency GWAS was likely underpowered in comparison to other traits tested, it was comparable in sample size to the lack of self-control GWAS. To the second point, these null findings parallel previous adolescent literature suggesting that urgency is more predictive of alcohol-related problems than measures of alcohol consumption, including binge drinking (Stautz & Cooper, 2013).

### Sample-Specific Findings

There are also several notable findings which did not replicate across samples but are nevertheless important to discuss. First, in the GENES sample, intoxication frequency slope was positively predicted by higher lack of self-control PGSs, an effect that was not observed in the other samples. The difference in PGS prediction of binge frequency (alcohol consumption PGS) versus intoxication frequency (lack of self-control PGS) slopes in GENES is striking given that measurements of binge and intoxication frequency were highly correlated across all timepoints (*r*>.80). This finding may speak to the importance of appreciating differences in even highly correlated phenotypes and the genetic influences which underlie them. More specifically, results may suggest differential influences on potentially nested outcomes where becoming intoxicated or “drunk (not just a little high) on alcohol,” endorsed by fewer participants at the highest levels of frequency, might reflect a correlated but higher intensity of drinking relative to binge drinking (i.e., 4+/5+ drink threshold; Levitt et al., 2009; Linden-Carmichael et al., 2021). In conjunction with support from extant phenotypic research (e.g., Bonar et al., 2022), we thus hypothesize that genetic risk underlying lack of self-control may be more relevant to high intensity or “extreme binge drinking.” In aggregate, results from the GENES sample demonstrate the nuanced but important influences of multiple genetic risk factors on binge drinking including both top-down (lack of self-control) and bottom-up (sensation seeking) constructs in a manner consistent with recent phenotypic studies conducted in adolescent and emerging adult samples, which suggest that both sensation seeking and lack of self-control are important predictors of changes in heavy episodic drinking (Ashenhurst et al., 2015; Ellingson et al., 2021; McCabe et al., 2021).

Second, the AUD PGSs predicted binge frequency growth model parameters *only* in ALSPAC and intoxication frequency growth model parameters *only* in Add Health. Specifically, AUD PGSs were associated with greater intercepts for both models (binge in ALSPAC and intoxication in Add Health), suggesting higher initial binge/intoxication drinking levels. AUD PGSs were also negatively associated with linear slope variance from T1-T4 (ages 16-20) for binge frequency in ALSPAC, similar to sensation seeking PGSs, such that those with higher PGSs displayed a more gradual slope or increase in binge frequency from age 16-20. This finding suggests that those with higher sensation seeking and AUD PGSs exhibit greater frequency of binge drinking in adolescence and that, while still persisting with greater than average frequency into emerging adulthood, the incremental increases are slower compared to the mean trajectory such that, by age 20, higher PGSs for these traits have seemingly little impact on binge frequency (max difference from mean level at age 20=|0.05| on scale of 1 to 4). In contrast, alcohol consumption PGSs continue to predict higher binge frequency relative to the mean into young adulthood. To date, there is limited support for these differential relations. For example, prior studies have suggested that genetic influences for alcohol consumption and AUD have distinct influences on changes in quantities of alcohol consumed (Deak et al., 2022) and further that AUD PGSs predict initial heavy drinking and early heavy drinking changes in adolescence (Li et al., 2017). Given the limited number of studies conducted in this area, the reported results provide an important contribution to this growing literature.

### Additional Considerations

In light of the numerous significant associations observed between PGSs and binge and intoxication frequency growth model parameters, two additional points merit discussion: (1) the effect sizes associated with studied PGSs and (2) noteworthy distinctions among the examined target samples. With respect to observed effect sizes, while individual PGSs were significantly associated with LGCM intercepts and slopes, the proportion of latent factor variance explained, after accounting for covariates, was relatively small even when estimating variance explained by two or more significant PGSs (i.e., 0.79-3.45% in Add Health and 2.10-3.49% in ALSPAC).

However, these estimates are comparable to those obtained in other cited PGS studies of substance use traits in developmental samples (e.g., Ksinan et al., 2019), and the impact on levels of binge and intoxication frequency may be significant in terms of the etiology and progression of normative drinking towards more problematic alcohol use. Higher PGSs often translated into meaningfully higher binge frequency categories at various timepoints, particularly at trajectory peak, compared to average and especially lower PGSs. For example, in ALSPAC a 1.5 *SD* increase in alcohol consumption PGS (87^th^ percentile) appeared to distinguish *monthly or less* from *monthly to weekly or more* binge drinking at developmental peak. Thus, while effect sizes are modest, prior research has shown that even subtle differences in binge drinking frequency may relate to greater odds of alcohol-related consequences and development of AUD (Dawson et al., 2005), highlighting the significance of the reported results.

It is also important to note differences in study characteristics that may have contributed to described differences in reported findings across studies. First, each study captures a slightly different developmental span. While Add Health and ALSPAC captured binge/intoxication frequency beginning in early-to-mid adolescence, recruitment and assessment for GENES, which followed a cohort of college students, began in late adolescence. Similarly, Add Health is unique in that assessments were conducted for participants much further into adulthood than either GENES or ALSPAC. Second, each study recruited participants from slightly different birth cohorts and employed different longitudinal designs and sampling strategies. Third, sample sizes, genotyping procedures, and phenotyping approaches (e.g., measures and assessment frequency) varied across studies. For example, the threshold for a ‘binge’ episode in ALSPAC is notably lower than the two U.S. samples when considering differences in definitions of standard drinks between the U.K. and the U.S. (8g of alcohol vs. 14g of alcohol; Kalinowski & Humphreys, 2016). While these differences bolster the observed evidence of consistent associations between sensation seeking and alcohol consumption PGSs and binge frequency growth parameters, they also highlight the potential importance of examining participant- and study-level characteristics which may obfuscate underlying genetic links.

### Limitations

This study is not without limitations which are worth noting as they provide points of opportunity and direction for future research. First, PGS were calculated and analyses conducted only in European ancestry samples primarily due to the absence of available large-scale GWAS of impulsive personality traits in non-European ancestry samples. This limits study generalizability and may ultimately contribute to health disparities if findings cannot be generalized to other ancestral populations, thus underscoring the importance of enhancing diversity in genetic studies (Martin et al., 2019). Second, the ability to detect potential associations between discovery GWAS summary statistics and target samples is based on the sample sizes and relative power of both. While sensation seeking and alcohol consumption PGSs showed the greatest prediction of binge and intoxication frequency changes in adolescence and young adulthood, in line with previous empirical studies and theory, these scores were also derived from the two most well-powered GWAS (Miller & Gizer, 2023, 2024). GWAS for lack of self-control and urgency were underpowered by comparison, and thus different patterns of associations may be detected in future studies as larger GWAS sample sizes become available for these traits (Sanchez-Roige et al., 2023).

## Conclusion

In sum, this study demonstrates the predictive utility of PGSs for dual-systems IPTs and alcohol use traits for longitudinal binge drinking phenotypes in three independent samples spanning adolescence and early adulthood. Across analyses conducted, samples examined, and phenotypes investigated, polygenic influences for sensation seeking and alcohol consumption emerged as reliable predictors of changes in binge drinking behaviors across adolescence and early adulthood with greater polygenic scores predicting increased binge drinking in adolescence and steeper drinking slopes across emerging adulthood. These findings are a successful extension of prior work demonstrating unique and additive genetic influences for sensation seeking and alcohol consumption and to a lesser extent lack of self-control and AUD on frequency of binge drinking and intoxication across relevant developmental time periods during which individuals often initiate binge drinking behaviors, increase quantity and frequency of binge drinking, and begin to experience problems related to excessive drinking.

## Data Availability

The full GWAS summary statistics for the 23andMe discovery data set will be made available through 23andMe to qualified researchers under an agreement with 23andMe that protects the privacy of the 23andMe participants. Please visit https://research.23andme.com/collaborate/#dataset-access/ for more information and to apply to access the data. GWAS summary statistics for risk taking in the UKB cohort along with ten smaller replication samples were obtained from https://thessgac.com/. GWAS summary statistics for drinks per week were obtained from https://conservancy.umn.edu/handle/11299/201564. Meta-analytic GWAS summary statistics (AlcGen, CHARGE +, and UKB) for grams of alcohol consumed per day were obtained through author request and the European Molecular Biology Laboratory’s European Bioinformatics Institute website (http://ftp.ebi.ac.uk/). PGC alcohol dependence and UKB GWAS summary statistics for AUDIT-C were obtained from the PGC website (https://www.med.unc.edu/pgc/). Million Veteran Program GWAS summary statistics were obtained through the Database for Genotypes and Phenotypes (dbGaP; Study Accession: phs001672). Finn-GenR6 ICD-based AUD GWAS data were obtained from https://r6.finngen.fi/pheno/AUD. For more information, visit https://finngen.gitbook.io/documentation/. Information on how to obtain Add Health data is available on the Add Health website (http://www.cpc.unc.edu/addhealth). The ALSPAC website contains details of all data that are available through a fully searchable data dictionary and variable search tool (http://www.bristol.ac.uk/alspac/researchers/our-data/).

## Acknowledgements

This research was supported by the National Institute on Alcohol Abuse and Alcoholism Grants F31AA027957 (principal investigator: Alex P. Miller), K01AA031724 (principal investigator: Alex P. Miller), F31AA029948 (principal investigator: Kellyn M. Spychala), R01AA013967 (principal investigator: Kim Fromme), and R01AA020637 (principal investigator: Kim Fromme); the National Institute on Drug Abuse Grant T32DA015035 (principal investigators: Kathleen K. Bucholz and Jeremy T. Goldbach); and a Center of Excellence award from the International Center for Responsible Gaming (principal investigator: Wendy S. Slutske). The authors have no known conflicts of interest to disclose. We would like to thank the research participants of 23andMe, Inc. for making this work possible. The full GWAS summary statistics for the 23andMe discovery data set will be made available through 23andMe to qualified researchers under an agreement with 23andMe that protects the privacy of the 23andMe participants. Please visit https://research.23andme.com/collaborate/#dataset-access/ for more information and to apply to access the data. GWAS summary statistics for risk taking in the UKB cohort along with ten smaller replication samples were obtained from https://thessgac.com/. GWAS summary statistics for drinks per week were obtained from https://conservancy.umn.edu/handle/11299/201564. Meta-analytic GWAS summary statistics (AlcGen, CHARGE +, and UKB) for grams of alcohol consumed per day were obtained through author request and the European Molecular Biology Laboratory’s European Bioinformatics Institute website (http://ftp.ebi.ac.uk/). PGC alcohol dependence and UKB GWAS summary statistics for AUDIT-C were obtained from the PGC website (https://www.med.unc.edu/pgc/).

Million Veteran Program GWAS summary statistics were obtained through the Database for Genotypes and Phenotypes (dbGaP; Study Accession: phs001672). FinnGenR6 ICD-based AUD GWAS data were obtained from https://r6.finngen.fi/pheno/AUD. For more information, visit https://finngen.gitbook.io/documentation/. This research uses data from Add Health, funded by grant P01 HD31921 (Harris) from the *Eunice Kennedy Shriver* National Institute of Child Health and Human Development (NICHD), with cooperative funding from 23 other federal agencies and foundations. Add Health is currently directed by Robert A. Hummer and funded by the National Institute on Aging cooperative agreements U01 AG071448 (Hummer) and U01AG071450 (Aiello and Hummer) at the University of North Carolina at Chapel Hill. Add Health was designed by J. Richard Udry, Peter S. Bearman, and Kathleen Mullan Harris at the University of North Carolina at Chapel Hill. No direct support was received from grant P01-HD31921 or cooperative agreements U01 AG071448 and U01AG071450. Information on how to obtain Add Health data is available on the Add Health website (http://www.cpc.unc.edu/addhealth). All procedures and described analyses utilizing the Genes and New Experiences Study (GENES) were approved by the University of Texas at Austin’s Institutional Review Board (Study: 2011-11-0042; Study Name: Mechanisms of Behavior Change). The UK Medical Research Council and Wellcome (Grant ref.: 217065/Z/19/Z) and the University of Bristol provide core support for ALSPAC. Genomewide genotyping data was generated by Sample Logistics and Genotyping Facilities at Wellcome Sanger Institute and LabCorp (Laboratory Corporation of America) using support from 23andMe. This publication is the work of the authors and Alex P. Miller, Ian R. Gizer, and Wendy S. Slutske will serve as guarantors for the contents of this paper. A comprehensive list of grants funding is available on the ALSPAC website. This research was specifically funded by the Wellcome Trust and Medical Research Council (MRC; Grant ref.: 092731, principal investigator: George Davey Smith), MRC and Alcohol research UK (Grant ref.: MR/L022206/1, principal investigator: Jon Heron), the National Institutes of Health (principal investigators: Matt Hickman, Glyn Lewis, and Ken Kendler; 5R01AA018333-05 and PD301198-SC101645, principal investigator: Matt Hickman), and MRC (Grant ref.: MR/L022206/1 and G0800612/86812, principal investigator: Matt Hickman). See also http://www.bristol.ac.uk/alspac/external/documents/grant-acknowledgements.pdf. We are extremely grateful to all the families who took part in this study, the midwives for their help in recruiting them and the whole ALSPAC team, which includes interviewers, computer and laboratory technicians, clerical workers, research scientists, volunteers, managers, receptionists and nurses. Computations for polygenic score generation were performed on the high-performance computing infrastructure provided by Research Computing Support Services and in part by the National Science Foundation under grant number CNS-1429294 at the University of Missouri, Columbia, MO. DOI: https://doi.org/10.32469/10355/69802 Model code is available upon request to Alex P. Miller.

This study was not preregistered. Alex P. Miller served as lead for conceptualization, formal analysis, methodology, visualization, and writing – original draft. Kellyn M. Spychala served in a supporting role for data curation. Wendy S. Slutske, Kim Fromme, and Ian R. Gizer served equally in supporting roles for conceptualization, data curation, investigation, funding acquisition, methodology, and resources. Wendy S. Slutske and Kim Fromme served equally in supporting roles for supervision, and Ian R. Gizer served as lead for supervision. All authors contributed equally to writing – review and editing.

