## Supplementary Methods for "Binge drinking trajectories across adolescence and early adulthood: Associations with genetic influences for dual-systems impulsive personality traits, alcohol consumption, and alcohol use disorder"

***Target Sample Genotyping, Imputation, and Quality Control***

**The National Longitudinal Study of Adolescent to Adult Health (Add Health).** At Wave 4, Oragene (Oragene™, DNAgenotek, Ottawa, Ontario, Canada) saliva samples were collected from consenting participants and DNA samples were genotyped using two Illumina (San Diego, CA) platforms: the Illumina Human Omni1-Quad BeadChip (∼1.1 million single nucleotide polymorphisms [SNPs] assayed genome-wide) and the Illumina Human Omni-2.5 Quad BeadChip (∼2.5 million SNPs assayed genome-wide). Raw genotype data were QCed by filtering out variants with a call rate < 90%, a minor allele frequency (MAF) < 0.5%, and ancestry-specific violations of Hardy-Weinberg equilibrium (HWE; *P* < 5 × 10^-5^). After QC procedures, genotype data were available for 9,974 individuals, and the Add Health genotype genome-wide association study (GWAS) data contained 609,130 SNPs common to both chips. Individuals of non-European ancestry (*n* = 4,187) were identified and removed from the sample using principal components analysis based on comparisons to ancestral principal components extracted from Phase 3 v5 1000 Genomes Project data (The 1000 Genomes Project Consortium, 2015).

Following initial QC, additional filters were applied prior to imputation of European ancestry genotypes (call rate < 98%, MAF < 1%, and HWE *P* < 5 × 10^-4^). The resulting QCed genotype data for individuals of European ancestry (*n* = 5,690; 346,754 SNPs) were imputed to Release 1 of the Haplotype Reference Consortium (HRCr1.1; McCarthy et al., 2016)). Imputation was conducted using the Michigan Imputation Server (Das et al., 2016; <https://imputationserver.sph.umich.edu>) and ShapeIt v2.r790 (Delaneau et al., 2013) following the GWAS and Sequencing Consortium of Alcohol and Nicotine Use (GSCAN) protocol (<https://genome.psych.umn.edu/index.php/GSCAN>) resulting in a final set of 38,953,236 high-quality genotyped and imputed variants. Principal components (PCs) indicating ancestry were calculated across individuals and HapMap3 reference samples using EIGENSTRAT (Price et al., 2006). See (Highland et al., 2018) and GSCAN website for additional genotype, quality control, and imputation details. Imputed genotype data and PCs for the Add Health sample were obtained through the Database for Genotypes and Phenotypes (dbGaP; Study Accession: phs001367).

For the current study, a genetic relatedness matrix (GRM) was also calculated in PLINK 1.9 (Chang et al., 2015), and using the steps outlined by Anderson et al. (2010), one individual from each pair with an identity by descent (IBD) *p̂* > 0.185 (i.e., second-degree relatives or closer) was removed from the sample (*n* = 462). Individuals self-reporting a race/ethnicity other than “White” were also excluded from the analytic sample (*n* = 160). Finally, PCs were used to identify ancestral outliers defined as any participant with a PC eigenvalue greater than or equal to four standard deviations from the mean on the first and/or second ancestry PC (i.e., the range present in European samples from Phase 3 v5 of the 1000 Genomes Project). Forty-seven participants met this exclusion criterion. Scatterplots of the principal component scores were examined to confirm that no ancestral outliers remained in the sample, and the first 10 PCs were used as covariates in longitudinal growth models described below to control for possible population stratification.

**Genes and New Experiences Study (GENES).** Participants provided Oragene (Oragene™, DNAgenotek, Ottawa, Ontario, Canada) saliva samples and extracted DNA samples were genotyped using the Illumina Infinium PsychArray BeadChip (San Diego, CA), which assays ∼265,000 SNPs across the genome. QC procedures on the non-Hispanic European ancestry GENES sub-sample (*n* = 376) included excluding variants and samples from statistical analyses based on call rate (< 98%), discrepant self-reported sex and biological sex, and relatedness (IBD *p̂* > 0.125; third-degree relatives or closer). Individuals self-reporting a race/ethnicity other than “White or Caucasian” were also excluded from the analytic sample (*n* = 11). Genotype data were imputed on the Michigan Imputation Server (Das et al., 2016; <https://imputationserver.sph.umich.edu>). Variants were phased with Eagle v2.3 (Loh et al., 2016) and imputed with Minimac3 1.0.13 (Das et al., 2016), using Phase 3 v5 of the 1000 Genomes Project (The 1000 Genomes Project Consortium, 2015) as a reference panel. MAF (< 1%) and HWE (*P* < 1 × 10^-5^) thresholds were applied to variants following phasing and imputation, yielding a final set of 5,250,123 high-quality genotyped and imputed variants (INFO > 0.9). Finally, flashPCA2 (Abraham et al., 2017) was used to calculate genomic ancestry PCs which were used to identify and remove ancestral outliers (± 4 *SD* on PC1 or PC2). Five participants met this exclusion criterion. Scatterplots of the principal component scores were examined to confirm that no ancestral outliers remained in the sample, and the first 10 PCs were used as covariates in longitudinal growth models described below to control for possible population stratification. See (Mallard et al., 2018) for additional details regarding genotyping, imputation, and quality control.

**Avon Longitudinal Study of Parents and Children (ALSPAC).** Pregnant women resident in Avon, UK with expected dates of delivery between 1st April 1991 and 31st December 1992 were invited to take part in the study. The initial number of pregnancies enrolled was 14,541 with 13,988 children alive at 1 year of age. When the oldest children were approximately 7 years of age, an attempt was made to bolster the initial sample with eligible cases who had failed to join the study originally. Thus, the total sample size for analyses using any data collected after the age of seven is 15,447 pregnancies and 14,901 children alive at 1 year of age. 14,203 unique mothers were initially enrolled in the study with additional recruitment providing a total of 14,833 unique women (G0 mothers). 12,113 G0 partners have been in contact with the study and 3,807 G0 partners are currently enrolled.

The ALSPAC offspring sample (G1) was genotyped from blood samples using the Illumina HumanHap550 Genotyping BeadChip (San Diego, CA), which assays ∼550,000 SNPs across the genome. Consent for biological samples has been collected in accordance with the Human Tissue Act (2004). The raw genotype data were then subjected to QC, including excluding variants with a call rate < 95%, a MAF < 1%, and violations of HWE (*P* < 5 × 10^-7^). All individuals of non-European ancestry were identified and removed from the analytic sample using multi-dimensional scaling analysis in comparison to the HapMap2 reference sample (The International HapMap Consortium, 2007). Offspring post-QC genotype data were then combined with those of the mothers and subjected to further QC, including excluding variants with a call rate of < 99% and HWE (*P* < 5 × 10^-7^). The genetic data were then imputed to HRCr1.1(McCarthy et al., 2016) using the Markov Chain Haplotyping algorithm (MaCH; Li et al., 2009). These procedures yielded a final set of 6,977,282 high-quality genotyped and imputed variants. Finally, a GRM and PCs indicating ancestry were calculated in PLINK 1.9 (Chang et al., 2015). The GRM was used to identify related individuals, and one individual from each pair with an IBD *p̂* > 0.185 (i.e., second-degree relatives or closer) was removed from the sample (*n* = 51). PCs were used to identify and remove ancestral outliers (± 4 *SD* on PC1 or PC2). Sixty-five participants met this exclusion criterion. Scatterplots of the principal component scores were examined to confirm that no ancestral outliers remained in the sample, and the first 10 PCs were used as covariates in longitudinal growth models described below to control for possible population stratification.

***Alternate Summary Statistics***

ALSPAC GWAS summary statistics for drinks per week (DPW) were included in the GSCAN meta-analytic sample (Liu et al., 2019) used in construction of the alcohol consumption GenomicSEM model from Miller & Gizer (2024). Thus, a secondary alcohol consumption GenomicSEM model was fit including only meta-analysis summary statistics for DPW from 23andMe (*N* = 403,939) and UK Biobank (UKB; *N* = 414,343) excluding ALSPAC and additional GSCAN samples. The genetic overlap between the full GSCAN DPW meta-analytic sample and the 23andMe + UKB drinks per week meta-analytic sub-sample (*N* = 818,282; $h_{g}^{2}$ = 0.048, *SE* = 0.002) was indistinguishable from 1 (*r*_g_ = 1.00, *SE* = 0.04, *P* = 3.31 × 10^-136^; see Supplementary Figure 1A for quantile-quantile (Q-Q) plot of secondary DPW meta-analysis). The resulting alcohol consumption common factor GWAS for the ALSPAC sample (*N_eff_* = 1,301,605) was similarly correlated with the summary statistics used to calculate the alcohol consumption polygenic scores (PGSs) in the other two samples (*r*_g_ = 1.00, *SE* = 0.03, *P* = 8.05 × 10^-185^; see Supplementary Figure 1B for Q-Q plot of secondary alcohol consumption GenomicSEM factor).

Similarly, the Add Health sample is included in the Psychiatric Genomics Consortium (PGC) alcohol dependence meta-analytic discovery sample (Walters et al., 2018) used in the construction of the larger alcohol use disorder (AUD) meta-analytic sample from Miller & Gizer (2024). Thus, a secondary AUD meta-analysis was conducted using a reduced set of PGC alcohol dependence summary statistics (*n_effective_* = 23,925) which included only unrelated genotyped individuals across samples and excluded Add Health summary statistic data. As expected, this alternate set of AUD summary statistics (*n_effective_* = 217,254; $h_{g}^{2}$ = 0.084, *SE* = 0.004) was perfectly correlated with the summary statistics derived in Miller & Gizer (2024; *r*_g_ = 1.04, *SE* = 0.05, *P* = 1.42 × 10^-96^; see Supplementary Figure 1C for Q-Q plot of secondary AUD meta-analysis).^[1]^ Moreover, the genetic correlation between the secondary alcohol consumption and AUD factors (*r*_g_ = .56, *SE* = 0.03, *P* = 7.55 × 10^-80^) was similar to that estimated from the primary summary statistics obtained in Miller & Gizer (2024; *r*_g_ = .58).

***PGS Methods***

Weights for PGSs were generated using SBayesR (Lloyd-Jones et al., 2019). SBayesR assumes that SNP effects are drawn from a finite mixture of normally-distributed priors and the key model parameters defining the genetic architecture of these SNPs are estimated through a Bayesian framework which shrinks SNP effect sizes while maximizing variance explained and accounting for linkage disequilibrium. Model parameters are randomly sampled following specification of default start values and iteratively updated through Markov Chain Monte Carlo (MCMC) methods which converge on high probability distributions of SNP effects. SBayesR has exhibited predictive performance comparable to other recently developed Bayesian PGS estimation methods (e.g., LDpred2, Privé et al., 2021; PRS-CS, Ge et al., 2019). These approaches may more accurately model SNP effects for psychiatric disorders and traits compared to *P-*value-based clumping and thresholding methods (Ni et al., 2021).

GWAS summary statistics were restricted to ∼1.1 million HapMap3 common variants (MAF>1%) excluding the major histocompatibility complex region due to its unusual linkage disequilibrium (LD) and genetic architecture (Finucane et al., 2015). These filtered SNPs were then matched to a sparse, banded LD matrix with a window size of 3 cM per SNP computed in a random sample of ∼50,000 unrelated individuals of European ancestry from UKB. Genome-wide SNP effects were estimated for each trait using a robust parameterization and 25,000 MCMC iterations with 5,000 burn-in samples and a thinning interval of 10. Individual PGSs were then calculated in PLINK 1.9 (Chang et al., 2015) using SBayesR-derived weights for participants in Add Health, GENES, and ALSPAC analytic samples.

Following specification of PGS conditional latent growth curve models, *R*^2^ change (Δ*R*^2^) was estimated for significant PGS predictors using an approach described by Hayes (2021). Provided analytic code, including an Δ*R*^2^ estimation function (*rsquareCalc*), from this publication was used to calculate the proportion of variance explained in intercept and slope factors by each significant PGS association after controlling for the other four PGSs as well as covariates: sex and the first 10 genomic ancestry principal components. Briefly, using this approach, Δ*R*^2^ is derived by direct matrix calculation (i.e., using sample correlations among model *x*’s and *y*) and subtraction of $R_{Reduced}^{2}$ from $R_{Full}^{2}$ where $R_{Full}^{2}$ is the proportion of latent factor variance explained by all model predictors and $R_{Reduced}^{2}$ is the proportion of latent factor variance explained by all model predictors with the target predictor or predictors omitted. In the case of more than one significant PGS predictor, the Δ*R*^2^ for inclusion of these PGSs was also estimated jointly.

***Binge Drinking and Intoxication Frequency Measurement***

**The National Longitudinal Study of Adolescent to Adult Health (Add Health).** The current study focused on unrelated Add Health participants of European ancestry who (1) completed one or more assessments of alcohol use across Waves 1 through 5 (ages 12 to 44) and (2) for whom there were also quality controlled (QC) genotype data available (see below for a description of genotyping and QC procedures). Binge drinking frequency was assessed at all five waves using the question: “During the past 12 months, on how many days did you drink {five = male/four = female} or more drinks in a row?” Intoxication frequency was assessed at Waves 1 through 4 using the question: “During the past 12 months, on how many days have you been drunk or very high on alcohol?” Because response options varied slightly across waves (i.e., 0 to 6 vs. 1 to 7) and the most extreme response options (“3 to 5 days a week”, “every day or almost every day”) were rarely endorsed, responses were binned and standardized across waves to reflect a 1 to 4 Likert scale (1=*Never*, 2=*Once per month or less*, 3=*2 or 3 days per month*, 4=*Weekly or more*).

**Genes and New Experiences Study (GENES).** The current study focused on GENES participants of European ancestry who (1) completed assessments of alcohol use at one or more timepoints across the course of the study and (2) for whom there were also quality controlled (QC) genotype data available (see below for a description of genotyping and QC procedures). Binge drinking frequency was captured at each assessment using the question: “During the last 3 months, how many times did you have {five = male/four = female} or more drinks at a sitting?” Likewise, intoxication frequency was captured at each assessment using the question: “During the last 3 months, how many times did you get drunk (not just a little high) on alcohol?” Frequencies were initially assessed using open-ended responses. Response data were significantly right-skewed and leptokurtic at each timepoint (binge drinking frequency: response ranges=0-30 to 0-90, skewness range=2.25-4.07, kurtosis range=5.04-23.97; intoxication frequency: response ranges=0-30 to 0-60, skewness range=2.52-3.65, kurtosis range=7.33-16.13). To eliminate bias related to violations of distributional assumptions of statistical analyses and to allow for greater comparison to the other two longitudinal samples, responses for each variable were binned and standardized across timepoints to reflect a 1 to 4 Likert scale (1=*Never*, 2=*Less than monthly*, 3=*Monthly*, 4=*Weekly or more*).

**Avon Longitudinal Study of Parents and Children (ALSPAC).** The current study focused on unrelated ALSPAC offspring of European ancestry who (1) completed assessments of alcohol use at one or more timepoints through mailed/online questionnaires and clinic visits (target ages = 16, 17.5, 18, 20, 22, 24, and 28 years old) and (2) for whom there were also QCed genotype data available (see below for a description of genotyping and QC procedures). ALSPAC data were collected and managed using REDCap electronic data capture tools hosted at the University of Bristol (Harris et al., 2009). REDCap (Research Electronic Data Capture) is a secure, web-based software platform designed to support data capture for research studies. At each assessment, binge drinking frequency was assessed using the question: “How often, during the past year, have you had six or more units [standard drinks] on one occasion?” As with Add Health, response options varied across timepoints (i.e., 1 to 5 vs. 1 to 6) and the most extreme response option (“Daily or almost daily”) was rarely endorsed. Thus, responses were similarly binned and standardized across timepoints to reflect a 1 to 4 Likert scale (1=*Never*, 2=*Less than monthly*, 3=*Monthly*, 4=*Weekly or more*). Informed consent for the use of data collected via questionnaires and clinics was obtained from participants following the recommendations of the ALSPAC Ethics and Law Committee at the time (<http://www.bristol.ac.uk/alspac/researchers/research-ethics/>).

***Latent Growth Curve Model Threshold Invariance***

Latent growth curve models (LGCMs) with ordinal variables assume threshold invariance across response options with respect to time (Mehta et al., 2004). That is, the relation between the underlying latent growth process and the measured response categories remains the same across assessment periods. We tested this assumption of threshold invariance as part of our modeling sequence by comparing relative fits of models in which thresholds were freed across age. Specifically, we first specified baseline models for all samples with no structure imposed on means, variances, or covariances of the underlying latent variable over time. Baseline models included only restrictions needed to scale the underlying latent variable (i.e., mean and variance of underlying variable set to 0 and 1 at T1, and first two thresholds of the underlying latent variables equal over time). These baseline models were then compared to models which additionally imposed equality of the third threshold for the underlying latent variable (i.e., equality of all thresholds over time) using scaled *χ*^2^differences tests for weighted least squares with mean and variance adjustment (Δ*χ*^2^_S_; Satorra, 2000). Constraining the third threshold did not result in worse fit than models constraining only the first two thresholds for any of the samples (*χ*^2^_S_ < 9.46, *df*=4-10, *P* > .092).

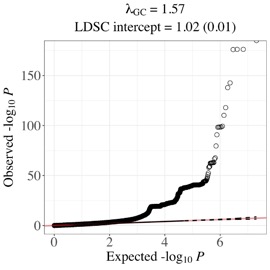

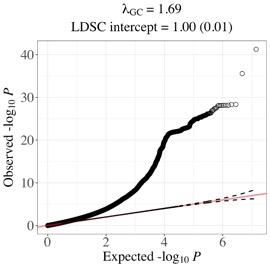

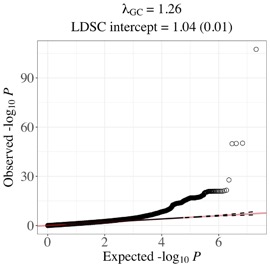

A.

B.

C.

**Supplementary Figure 1. Q-Q plots for secondary GWAS conducted for ALSPAC and Add**

**Health alcohol consumption and AUD PGS calculations.** These results have not

been adjusted for genomic control inflation factors (λGC). **(A)** 23andMe + UK

Biobank drinks per week meta-analysis. **(B)** Secondary GenomicSEM alcohol

consumption common factor GWAS. **(C)** Secondary AUD GWAS meta-analysis.

1. Note that because LDSC is not a bounded estimator, it can produce estimates outside of [-1, 1] due to sampling variation (Bulik-Sullivan et al., 2015).
